# Impact of immune heterogeneity on antigenic evolution

**DOI:** 10.64898/2026.11.22.26368671

**Authors:** Myrthe S Willemsen, Richard A Neher, Ganna Rozhnova

**Affiliations:** Julius Center for Health Sciences and Primary Care, University Medical Center Utrecht, Utrecht University, Utrecht, The Netherlands; Center for Complex Systems Studies (CCSS), Utrecht University, Utrecht, The Netherlands; Biozentrum, University of Basel, Basel, Switzerland; Swiss Institute of Bioinformatics, Basel, Switzerland; BioISI—Biosystems & Integrative Sciences Institute, Faculty of Sciences, University of Lisbon, Lisbon, Portugal; Faculty of Sciences, University of Lisbon, Lisbon, Portugal

**Author notes:** **Author for correspondence:** Myrthe S Willemsen.

**Keywords:** Antigenic evolution, Cross-immunity, Viruses, Immune heterogeneity, Age

## Abstract

Antigenic evolution allows pathogens to escape pre-existing host immunity, but hosts differ in their immune histories because of population structure, demographic processes, and stochastic variation. Whether such immune heterogeneity promotes pathogen antigenic diversification remains poorly understood. We investigated how different sources of immune heterogeneity affect antigenic evolution. We developed a status-based multi-strain transmission model in which novel strains emerge stochastically within a tree-structured antigenic space. We examined immune heterogeneity arising from mixing between host subpopulations, aging, and stochastic variation in immune profiles, and evaluated its effects on strain coexistence and phylogenetic diversity. Limited mixing generated immune differences between host subpopulations, but increased phylogenetic diversity only under strong population separation. Aging alone had little effect on evolutionary dynamics but enhanced the effect of limited mixing on phylogenetic diversity. Stochastic immune variation promoted strain replacement rather than coexistence, primarily through variation in the overall susceptible population available to newly emerging strains. In general, immune heterogeneity alone does not necessarily promote antigenic diversification. Its effects depend on how immune differences are generated and maintained within the host population. More broadly, our findings highlight the importance of understanding how assumptions linking epidemiological and evolutionary processes shape the emergent dynamics of evolutionary-epidemiological models.

## Introduction

Antigenic evolution allows many pathogens to escape pre-existing host immunity and persist in host populations. Rapid antigenic change has been observed in pathogens including respiratory syncytial virus [1], SARS-CoV-2 [2], and seasonal influenza [3, 4]. Because host immunity determines which strains can spread successfully, heterogeneity in immunity across a host population may alter the selective pressures acting on pathogen strains. In particular, if subpopulations of hosts differ in their susceptibility to antigenically distinct strains, these subpopulations may provide distinct immunological niches in which different strains can circulate. Such strain-specific immune heterogeneity could therefore affect strain competition and, over time, antigenic evolution.

Several mechanisms can generate heterogeneity in immune histories. When mixing between host subpopulations is limited because of geographical or social separation, different strains may circulate within different subpopulations. Their epidemic and immune histories can consequently diverge, potentially reinforcing differences in susceptibility to subsequently emerging strains. Geographical variation in strain circulation is well documented for seasonal influenza [5, 6] and has also been observed for SARS-CoV-2 [7]. Immune histories may also differ systematically between age groups because exposure to infection and vaccination accumulates with age. For influenza, neutralization assays have shown that immunity and the effects of viral mutations differ between age groups, suggesting that age groups may exert different selection pressures on circulating viruses [8]. Finally, random variation in immune profiles may arise from differences in HLA types [9], vaccination histories, seasonality, immune imprinting [10–12], and other sources of variation.

Despite these multiple sources of immune heterogeneity, their consequences for long-term antigenic evolution remain poorly understood. Previous modeling studies have investigated how mixing and host heterogeneity, including differences in immunity, affect particular evolutionary processes such as strain emergence [13, 14], coexistence between two competing strains [15], or overall disease persistence [16–18]. However, it remains unclear whether and under what conditions immune heterogeneity alters pathogen diversification when novel strains continuously arise and compete in a partially immune host population. Understanding this could ultimately help inform targeted interventions in populations that contribute disproportionately to the emergence of novel strains.

We hypothesize that strain-specific differences in immunity can reduce competition between pathogen strains by creating distinct subpopulations of susceptible hosts. Different strains may then preferentially circulate within these host subpopulations and independently generate antigenically novel descendants. Over time, this process could promote strain coexistence and increase phylogenetic diversity at the population level. Importantly, this mechanism may depend on how immune heterogeneity is generated and maintained. Structured differences in immune histories between host subpopulations may have different evolutionary consequences from stochastic variation in immune profiles.

Here, we investigate whether, and under what conditions, immune heterogeneity shapes pathogen antigenic evolution. We developed a parsimonious multi-strain model in which pathogen strains compete for susceptible hosts and interact through partial cross-immunity induced by previous infections. The model builds on a compartmental status-based framework that tracks the proportions of individuals susceptible to different strains [19–22], which allows immune heterogeneity to be represented while remaining computationally tractable over long evolutionary timescales. We extended this framework by introducing novel strains stochastically within a tree-structured antigenic space. Using this model, we examine three mechanisms that can generate immune heterogeneity, i.e., mixing between host subpopulations, aging, and stochastic variation in immune profiles, and determine how each affects strain coexistence and antigenic diversity. Finally, we examine how assumptions governing the emergence of novel strains influence these evolutionary outcomes.

## Methods

### Overview

We developed a multi-strain compartmental model to investigate how immune differences between host subpopulations affect antigenic evolution (hereafter, ‘evolution’). The model combines deterministic epidemiological dynamics with the stochastic emergence of novel strains in a tree-structured antigenic space. The host population is divided into multiple subpopulations connected through a contact matrix, with pathogen strains interacting through cross-immunity generated by previous infections. We considered three mechanisms that can generate immune differences between subpopulations: mixing, aging, and stochastic immune variation. We quantified the resulting epidemiological differences between subpopulations and their effects on two aspects of antigenic evolution: strain coexistence and phylogenetic diversity.

### Evolutionary-epidemiological model

#### Population structure and transmission

We used a status-based multi-strain model [12, 23, 24], previously extended to multiple host subpopulations [22].

The model describes the circulation of *N* pathogen strains in a host population divided into *M* subpopulations. Unless otherwise specified, subpopulations are assumed to be of equal and constant size.

For each strain *a* = 1, … , *N* and subpopulation 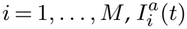 denotes the proportion of individuals infected with strain *a* at time *t*, and 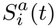 the proportion susceptible to strain *a* at time *t*. These are strain-specific states rather than mutually exclusive population compartments. Susceptibility and infection status are tracked separately for each strain, while immunity is represented implicitly through the corresponding reduction in 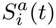 due to infection. The model allows for coinfection with different strains, but co-infecting strains do not interact directly within hosts, i.e., interactions between strains occur only indirectly through cross-immunity. The epidemiological dynamics are described by

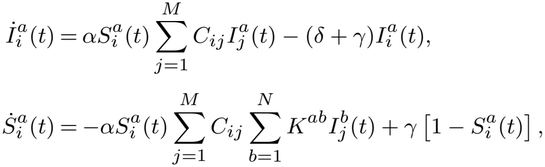

where *i* = 1, … , *M* , *a* = 1, … , *N* and the parameters are defined in Table 1. A schematic of the model is shown in Supplement Figure 5.

**Table 1.** Model parameters and variables. Unless otherwise specified, simulations used the default parameter values listed here.

| **Symbol** | **Parameter definition** (unit) | **Default value** | **Ranges tested** |
|---|---|---|---|
| *N* | Maximum number of co-circulating strains | 25 | (sensitivity analysis: 5 to 40) |
| *M* | Number of subpopulations | 10 | – |
| α | Transmission rate; see [23] (*t*<sup>−1</sup>) | 2 | – |
| γ | Rate of population turnover (*t*<sup>−1</sup>) | 10<sup>−4</sup> | – |
| δ | Recovery rate (*t*<sup>−1</sup>) | 1 | – |
| σ | Magnitude of stochastic immune variation around the expected susceptibility | 0 | 0 to 1 |
| *d* | Cross-immunity breadth (units of antigenic mutations) | 10 | (sensitivity analysis: 1, 100) |
| *t*<sub>e</sub> | Mean of the exponential distribution of times between strain-emergence events (*t*) | 10 | (sensitivity analysis: 100) |
| *C*<sub>*ij*</sub> | Proportion of all contacts made by an individual in group *i* that occur with individuals in group *j* | – | – |
| *c* | Mixing parameter defined as the ratio of between- to within-subpopulation contacts, $c = \frac{C_{ij}}{C_{ii}}, \qquad i \neq j$ | 1 | 1 to 1 · 10<sup>−5</sup> |
| *K*<sup>*ab*</sup> | Probability that exposure to strain *b* confers immunity to strain *a* | – | – |
| *S*<sub>*i*</sub><sup>*a*</sup> | Proportion of individuals susceptible to strain *a* in subpopulation *i* | – | – |
| *I*<sub>*i*</sub><sup>*a*</sup> | Proportion of individuals infected with strain *a* in population *i* | 10<sup>−6</sup> at emergence | – |
| *a*, *b* = 1, . . . , *N* | Indices used to denote strains | – | – |
| *i*, *j* = 1, . . . , *M* | Indices used to denote subpopulations | – | – |

Interactions between susceptible and infected individuals lead to new infections. Infections in a given subpopulation arise from interactions with infected individuals across all subpopulations. Transmission of strain *a* to susceptible individuals in subpopulation *i* occurs at a total rate

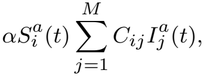

where *α* is the transmission rate and *C*_*ij*_ is the proportion of all contacts made by an individual in subpopulation *i* with individuals in subpopulation *j*. We assumed a symmetric contact matrix, *C*_*ij*_ = *C*_*ji*_, normalized such that 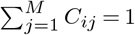 for each subpopulation *i*. Infected individuals recover at rate *δ*, while population turnover due to birth and death occurs at rate *γ*. Note that individuals immune to strain *a* are not represented explicitly, and therefore 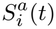 and 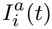 do not partition the population, i.e.,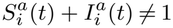.

Previous infections reduce susceptibility to antigenically related strains through the cross-immunity term *K*^*ab*^. Thus, exposure to individuals infected with strain *b* reduces the proportion susceptible to strain *a* at a rate determined by the antigenic distance between the two strains. Cross-immunity and its dependence on antigenic distance are defined below.

#### Cross-immunity and antigenic distance

We defined *K*^*ab*^ as the probability that exposure to strain *b* confers immunity to strain *a*, with 0 *≤ K*^*ab*^ *≤* 1. We assumed complete homologous immunity, such that *K*^*aa*^ = 1, and cross-immunity between different strains decreased exponentially with their antigenic distance [10, 12, 23, 24]:

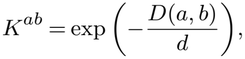

where *D*(*a, b*) is the antigenic distance between strains *a* and *b*, and *d* determines the breadth of cross-immunity. Larger values of *d* correspond to broader cross-immunity, such that protection decays more slowly with antigenic distance. Smaller values correspond to more strain-specific immunity. Accordingly, *K*^*ab*^ = 1 represents complete protection, whereas values approaching 0 represent increasingly weak cross-immunity.

Strains occupy a tree-structured antigenic space. Each newly emerging strain differs from its parent by one antigenic mutation and therefore has an antigenic distance of one from its parent. The antigenic distance *D*(*a, b*) between any two strains is defined as the number of mutations along the unique path connecting them on the phylogenetic tree. Thus, *D*(*a, a*) = 0, directly related strains separated by one mutation have *D*(*a, b*) = 1, and increasingly antigenically distant strains have progressively larger values of *D*(*a, b*). We assumed no recurrent or back mutations. For a novel strain *a*, its parent *p* lies on the path to any other strain *b*, allowing their distance to be computed as *D*(*a, b*) = *D*(*p, b*) + 1.

#### Strain emergence and extinction

Novel strains emerged at stochastic time intervals drawn from an exponential distribution with mean *t*_*e*_. At each emergence event, a parent strain and the subpopulation in which its descendant emerged were jointly selected from the currently circulating strains and subpopulations, respectively. Each novel strain differed from its parent by one antigenic mutation and was introduced at an initial prevalence of 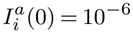 in the selected subpopulation *i*.

We considered two mechanisms linking strain emergence to the epidemiological dynamics. In the uniform emergence mechanism, used for the main analyses, the parent strain and subpopulation were jointly selected uniformly at random.

Under this mechanism, the opportunity to generate an antigenically novel descendant is independent of the current prevalence of a strain and of its prevalence within a particular subpopulation. Uniform emergence therefore separates the generation of antigenic novelty from the epidemiological success of circulating strains.

We additionally considered a prevalence-weighted emergence mechanism, in which the probability that a novel strain descended from strain *a* in subpopulation *i* was proportional to its current prevalence,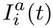. Under this mechanism, strains causing more infections have proportionally more opportunities to generate antigenically novel descendants. Prevalence-weighted emergence therefore couples the generation of antigenic novelty to the epidemiological abundance of circulating strains. Comparing the two mechanisms allowed us to determine whether the effects of immune differences on evolutionary dynamics depended on this coupling between epidemiological success and the generation of antigenic novelty.

When a novel strain *a* emerged, part of the host population was already immune through previous exposure to antigenically related strains. However, the proportion of individuals susceptible to this strain in subpopulation *i*, 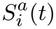, had not been tracked. Calculating 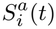exactly would require reconstructing the effects of all previous infections, which would be computationally demanding. We therefore derived an approximation for 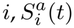 based on the proportion of individuals susceptible to its parent strain *p* as

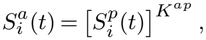

where *K*^*ap*^ is the cross-immunity between the novel strain *a* and its parent *p*. A derivation of this approximation is provided in Supplementary Section (b).

While novel strains emerged, circulating strains could also become extinct. To keep the number of simultaneously represented strains computationally tractable, we allowed at most *N* ^max^ co-circulating strains. At each strain-emergence event, strains with a prevalence below 10^*−*7^ were considered extinct and removed from the system. If the number of circulating strains had reached *N* ^max^ and no strain was below this threshold, we removed the least prevalent strain with a negative growth rate. If no circulating strain had a negative growth rate, the least prevalent strain was removed. The complete procedure is described in Algorithm 1. Strain extinction was therefore evaluated at strain-emergence events rather than occurring stochastically between these events.

### Mechanisms generating immune differences between subpopulations

We considered three mechanisms that can generate immune differences between host subpopulations: mixing, aging, and stochastic immune variation. These mechanisms represent distinct ways in which strain-specific susceptibility profiles can become heterogeneous across a host population. Limited mixing allows subpopulations to develop different epidemiological and immune histories through partially separated transmission dynamics. Aging generates systematic differences in accumulated immunity between age groups. Stochastic immune variation introduces random differences in susceptibility to newly emerging strains. We examined each mechanism separately and additionally considered the combined effects of aging and limited mixing.

#### Mixing

We first considered whether limited mixing could generate immune differences by allowing pathogen strains to circulate differently across otherwise identical host subpopulations. Mixing was controlled through the ratio of between-to within-subpopulation contact rates,

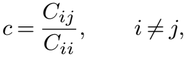

where all within-subpopulation contact rates are assumed to be equal (*C*_*ii*_ = *C*_*jj*_) and all between-subpopulation contact rates are assumed to be equal (*C*_*ij*_ = *C*_*kl*_ for all *i* ≠ *j* and *k≠*= *l*). Because each row of the contact matrix was normalized such that 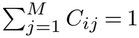 , the contact rates were given by

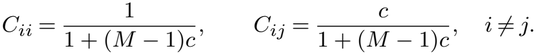

At *c* = 1, contacts occur equally within and between subpopulations, corresponding to homogeneous mixing. As *c* decreases below 1, contacts become increasingly concentrated within subpopulations, and transmission between subpopulations becomes progressively more limited. In the limit *c →* 0, subpopulations become epidemiologically isolated. We varied *c* from 1 to 1 *·* 10^*−*5^ in the main analyses, with smaller values representing stronger population separation.

Mixing does not impose immune differences directly. Instead, it can generate them endogenously because reduced transmission between subpopulations can cause strains to circulate at different frequencies across subpopulations, leading their accumulated strain-specific susceptibility profiles to diverge over time. We therefore examined whether increasing population separation generated differences in strain circulation and immunity and whether these differences were accompanied by changes in strain coexistence and phylogenetic diversity.

#### Aging

We next considered aging as a mechanism that can generate systematic differences in immunity between host subpopulations. In this scenario, the *M* subpopulations represent consecutive age groups, with *i* = 1 denoting the youngest and *i* = *M* the oldest age group. Fully susceptible individuals enter the youngest age group and subsequently progress through successive age groups. As a result, older age groups have had more time to accumulate immunity through previous infections, whereas the continuous entry of susceptible individuals into the youngest age group can generate differences in susceptibility between age groups.

To maintain equal-sized age groups and a constant total population size, individuals progress from age group *i* to age group (*i* + 1) at rate *γM* , with the same rate determining entry into the youngest age group and removal from the oldest. The epidemiological dynamics are therefore given by

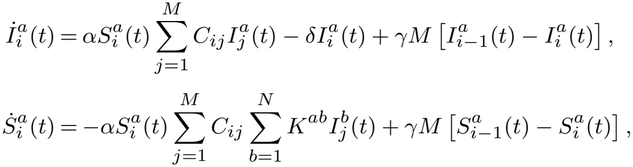

where *i* = 1, … , *M* , *a* = 1, … , *N*. The virtual compartments 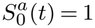 and 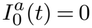 represent the entry of newborn individuals who are fully susceptible and uninfected. In this scenario, progression through age groups replaces the population-turnover terms used in the model without aging. The expected duration in each age group is therefore 1*/*(*γM*), giving an expected lifespan of 1*/γ*.

In the analyses, we first considered aging under homogeneous mixing (*c* = 1) to isolate the effect of age-related differences in accumulated immunity from differences generated by population separation. We subsequently varied *c* from 1 to 1 *·* 10^*−*5^ to examine whether aging modified the effects of mixing on epidemiological and evolutionary dynamics.

#### Stochastic immune variation

Finally, we considered stochastic immune variation as a mechanism generating immune differences between subpopulations. For each newly emerging strain *a*, we first calculated the expected susceptibility of subpopulation *i*, 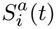, from its susceptibility to the parent strain *p*, as described in Section ‘Strain emergence and extinction’. We then introduced stochastic variation around this expected susceptibility by independently sampling the susceptibility 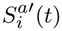 for each subpopulation from a beta distribution,

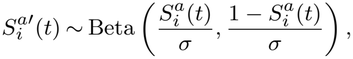

where *σ* controls the magnitude of stochastic variation around the expected susceptibility. Under this parameterization,

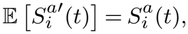

while its variance is

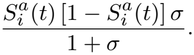

Thus, as *σ →* 0, the variance approaches zero and the sampled susceptibility 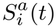 converges to the expected susceptibility 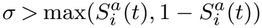. For *σ >* 0, the sampled susceptibility can differ from its expected value, with larger values of *σ* generating greater variation. For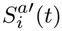, the beta distribution becomes bimodal, concentrating the sampled susceptibility values near 0 and 1. We varied *σ* from 0 to 0.1, ensuring that the beta distributions remained unimodal. Because susceptibilities are sampled independently for each subpopulation, increasing *σ* can generate increasing immune differences between subpopulations.

As a consequence of this formulation, stochastic immune variation can also change the total susceptible population available to a newly emerging strain. In particular, the total sampled susceptibility, 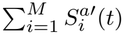, is not constrained to equal the expected total susceptibility,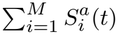. Some emerging strains can therefore encounter a larger susceptible population than expected, whereas others encounter a smaller one. Thus, stochastic immune variation simultaneously generates immune differences between subpopulations and variation among emerging strains in the overall susceptible population available to them.

To distinguish these two effects, we considered a control scenario in which variation in the overall susceptibility to newly emerging strains was retained without introducing additional immune differences between subpopulations. We first sampled 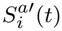 independently for each subpopulation as described above. We then rescaled the expected susceptibilities according to

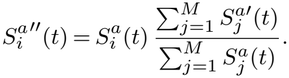

Because the same scaling factor is applied to all subpopulations, this procedure preserves their relative expected susceptibility profile while ensuring that the total susceptibility is identical to that obtained from the stochastic draws,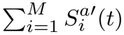. The control scenario therefore retains variation among emerging strains in the overall susceptible population available to them without introducing the additional immune differences between subpopulations generated by independent stochastic sampling. Comparing the two scenarios allowed us to distinguish the effects of stochastic immune differences between subpopulations from those arising from variation in the overall susceptible population available to newly emerging strains.

### Outcome measures

We quantified both epidemiological and evolutionary outcomes to determine how the different mechanisms generating immune differences affected strain circulation, immunity, and antigenic evolution. Epidemiological outcomes characterized infection dynamics and differences between host subpopulations, whereas evolutionary outcomes characterized the coexistence and phylogenetic relationships of circulating strains.

#### Epidemiological outcomes

##### Infection prevalence

We quantified the total infection prevalence within subpopulation *i* as

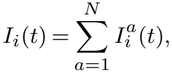

and the average infection prevalence across the *M* equally sized subpopulations as

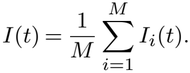

Because infection states are defined separately for each strain and coinfection is permitted, *I*_*i*_(*t*) and *I*(*t*) represent the mean number of concurrent strain-specific infections per individual rather than the proportion of individuals infected with at least one strain. Their theoretical range is therefore [0, *N*]. A value of 0 indicates no infection, whereas values greater than 1 are possible when individuals are infected with multiple strains.

##### Strain frequencies

We calculated the frequency of each strain among all strain-specific infections with subpopulation *i*,

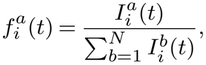

and the strain frequencies among all subpopulations,

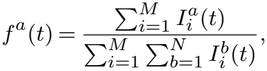

such that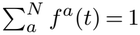.

##### Exposure originating from other subpopulations

To quantify the degree to which infection dynamics in a subpopulation were driven by transmission from outside that subpopulation, we calculated the proportion of infectious exposure experienced by individuals in subpopulation *i* that originated from other subpopulations:

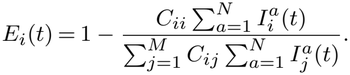

This measure ranges from 0 to 1. A value of 0 indicates that all infectious exposure originates within the focal subpopulation, whereas a value approaching 1 indicates that most exposure originates from other subpopulations. We averaged this measure across subpopulations to quantify between-subpopulation exposure at the population level.

##### Differences in strain circulation between subpopulations

To determine whether different strains circulated in different subpopulations, we compared their strain-specific infection profiles. For each pair of subpopulations *i* and *j*, we calculated the cosine distance between the vectors of strain-specific infections,

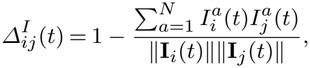

where 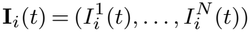 and ∥**I**_*i*_(*t*)∥ denotes its Euclidean norm. We then averaged pairwise distances across subpopulations,

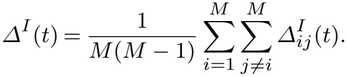

Because all components of the infection profiles are non-negative, *Δ*^*I*^ (*t*) ranges from 0 to 1. A value of 0 indicates that subpopulations have identical relative strain-circulation profiles, whereas larger values indicate increasingly different distributions of circulating strains across subpopulations. The measure is insensitive to differences in the overall magnitude of infection prevalence and therefore specifically captures differences in the relative composition of circulating strains.

##### Immune differences between subpopulations

We similarly quantified immune differences between subpopulations from their strain-specific susceptibility profiles. For each pair of subpopulations *i* and *j*, we calculated

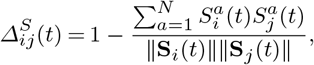

Where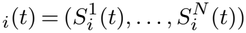. Pairwise distances were averaged across subpopulations,

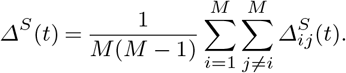

As for differences in strain circulation, *Δ*^*S*^ (*t*) ranges from 0 to 1. A value of 0 indicates identical relative strain-specific susceptibility profiles across subpopulations, whereas larger values indicate greater strain-specific immune differences. Because cosine distance depends on the relative pattern of susceptibility across strains rather than its overall magnitude, this measure distinguishes strain-specific immune differences from uniform differences in overall susceptibility between subpopulations.

#### Evolutionary outcomes

We characterized two complementary aspects of the evolutionary dynamics. First, strain coexistence measures how many strains circulate simultaneously and how evenly infections are distributed among them. Second, phylogenetic diversity measures how evolutionarily separated the circulating strains are. These measures distinguish the coexistence of multiple closely related strains from the persistence of divergent evolutionary lineages.

##### Strain coexistence

We quantified strain coexistence using the Shannon entropy of the strain frequencies,

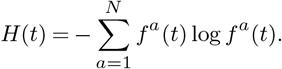

For *N* circulating strains, *H*(*t*) ranges from 0 to log *N*. It equals 0 when a single strain accounts for all infections and increases as more strains circulate and their frequencies become more evenly distributed. The maximum, log *N* , occurs when all *N* strains circulate at equal frequencies. Shannon entropy therefore captures both the number of co-circulating strains and the evenness of their relative frequencies.

##### Phylogenetic diversity

Strain coexistence alone does not indicate whether co-circulating strains belong to closely related or divergent evolutionary lineages. We therefore additionally quantified phylogenetic diversity using a frequency-weighted average pairwise measure based on the time to the most recent common ancestor (*T*_MRCA_) of circulating strains:

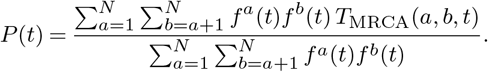

Here, *T*_MRCA_(*a, b, t*) denotes time elapsed since the most recent common ancestor of strains *a* and *b* emerged. If one strain was an ancestor of the other, that strain constituted their most recent common ancestor.

The weighting by *f*^*a*^(*t*)*f*^*b*^(*t*) gives greater influence to pairs of strains that account for a larger proportion of current infections. A value approaching 0 indicates that the dominant circulating strains share very recent common ancestry, whereas larger values indicate the simultaneous circulation of lineages that have remained evolutionarily separated for longer. This is reflected by increased branching in the phylogenetic tree, with individual strains giving rise to more descendant lineages. We used the average pairwise *T*_MRCA_ rather than the *T*_MRCA_ of all circulating strains because the latter is highly sensitive to the extinction of individual lineages. The pairwise measure is less sensitive to such events and provides a continuous measure of phylogenetic diversification.

##### Descendant frequency and lineage fixation

To visualize the persistence and replacement of evolutionary lineages, we additionally calculated the descendant frequency of strain *a*,

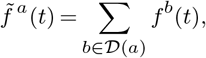

where *D*(*a*) contains strain *a* and all of its descendants. Descendant frequency ranges from 0 to 1 and represents the fraction of current infections belonging to the lineage founded by strain *a*. Values approaching 1 indicate that this lineage accounts for almost all circulating infections and therefore approaches lineage fixation, whereas values approaching 0 indicate that the lineage has largely disappeared from the circulating pathogen population.

### Simulation and implementation

We initialized each simulation with a single strain. Initial susceptibilities were set to *δ/α*, which is the equilibrium value. Each simulation was run for 3,000 *δ*^*−*1^ and outcomes were averaged over time. For each parameter set, we report the median and interdecile range over 100 stochastic simulations. Unless otherwise specified, simulations used the parameter values given in Table 1.

The model and simulations were implemented in Python version 3.13.2. The code is available at https://github.com/MyrtheW/immune_heterogeneity_evolution. Details of the model implementation and required packages are provided in Supplementary Section (a).

### Sensitivity analyses

We assessed the robustness of the evolutionary dynamics to two assumptions governing the emergence and persistence of strains: the frequency of strain emergence and the maximum number of co-circulating strains. First, we varied the

mean time between strain-emergence events, *t*_*e*_, to determine whether the frequency at which novel strains entered the system affected the evolutionary dynamics. The main analyses used *t*_*e*_ = 10 *δ*^*−*1^, and we compared these results with less frequent strain emergence, *t*_*e*_ = 100 *δ*^*−*1^. Second, we varied the maximum number of co-circulating strains, *N* , to determine whether the imposed computational limit constrained the evolutionary dynamics. We used *N* = 25 in the main analyses because increasing this limit further had negligible effects on the outcome measures across the parameter ranges considered. Throughout the manuscript, time is expressed in units of *δ*^*−*1^.

## Results

### Baseline epidemiological and evolutionary dynamics

We first considered the model dynamics in the absence of immune differences between subpopulations. Under homogeneous mixing, newly emerging strains spread rapidly across subpopulations and generated recurrent changes in strain frequencies, accompanied by fluctuations in overall infection prevalence (Figure 1). Over time, strains were replaced or gave rise to co-circulating lineages, resulting in branching of the phylogenetic tree. The balance between strain replacement and phylogenetic branching depended on the breadth of cross-immunity (Supplement Figure 6).

**Figure 1.**
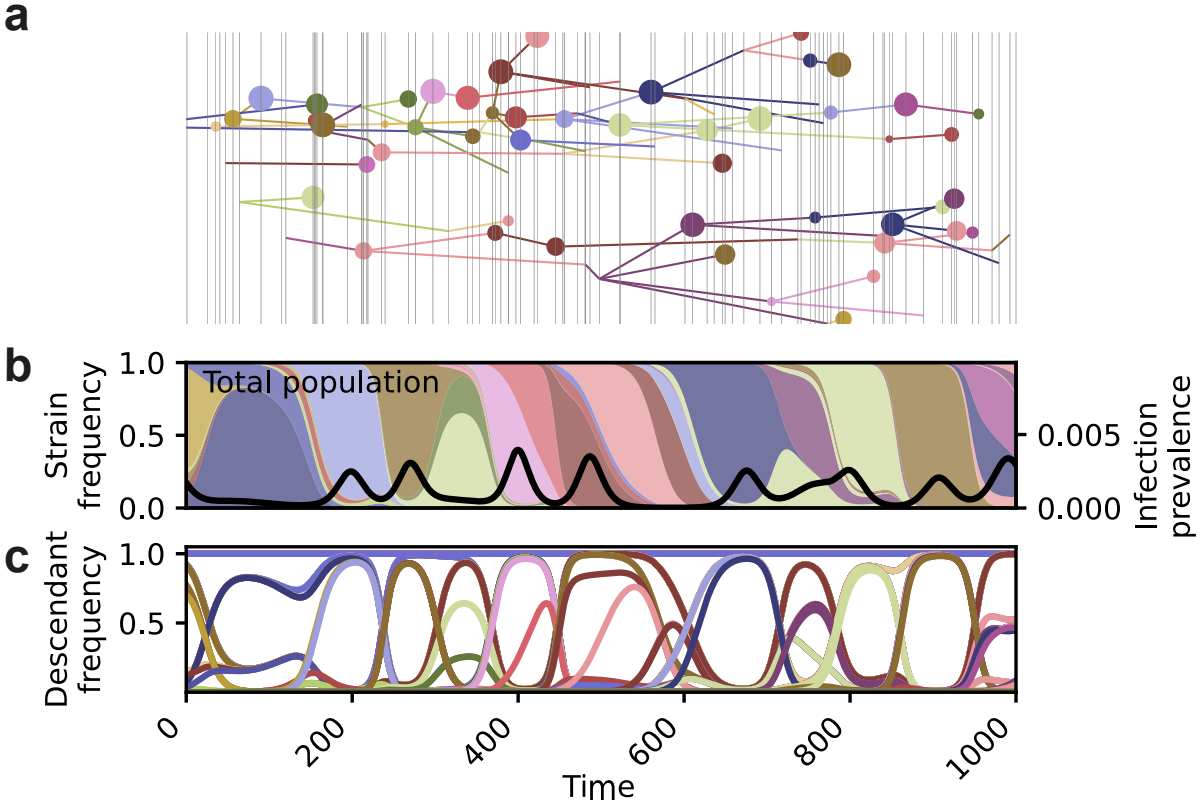
Evolutionary and epidemiological dynamics of the baseline model in the absence of immune differences between subpopulations. Model dynamics over a period of 1000 *δ*^*−*1^ under homogeneous mixing and baseline parameter values (Table 1). **a** Phylogenetic tree of circulating strains. The horizontal position of each node indicates the time at which the corresponding strain emerged, while the vertical position is arbitrary. Light grey vertical lines indicate strain-introduction events, and node size is proportional to the cumulative prevalence of the corresponding strain. **b** Relative frequencies of circulating strains (colored areas, left y-axis) and overall infection prevalence (black line, right y-axis). **c** Frequency of each strain together with all of its descendants, illustrating the persistence and fixation of individual lineages. Colors identify the same strains across panels.

### Limited mixing affects evolution only under strong population separation

We first investigated how mixing between subpopulations affected strain circulation, immune differences, and evolutionary dynamics (Figure 2). Under homogeneous mixing (*c* = 1), individuals were equally likely to contact individuals within their own or other subpopulations. As mixing decreased, contacts became increasingly concentrated within subpopulations, reducing exposure to infections originating from other subpopulations.

**Figure 2.**
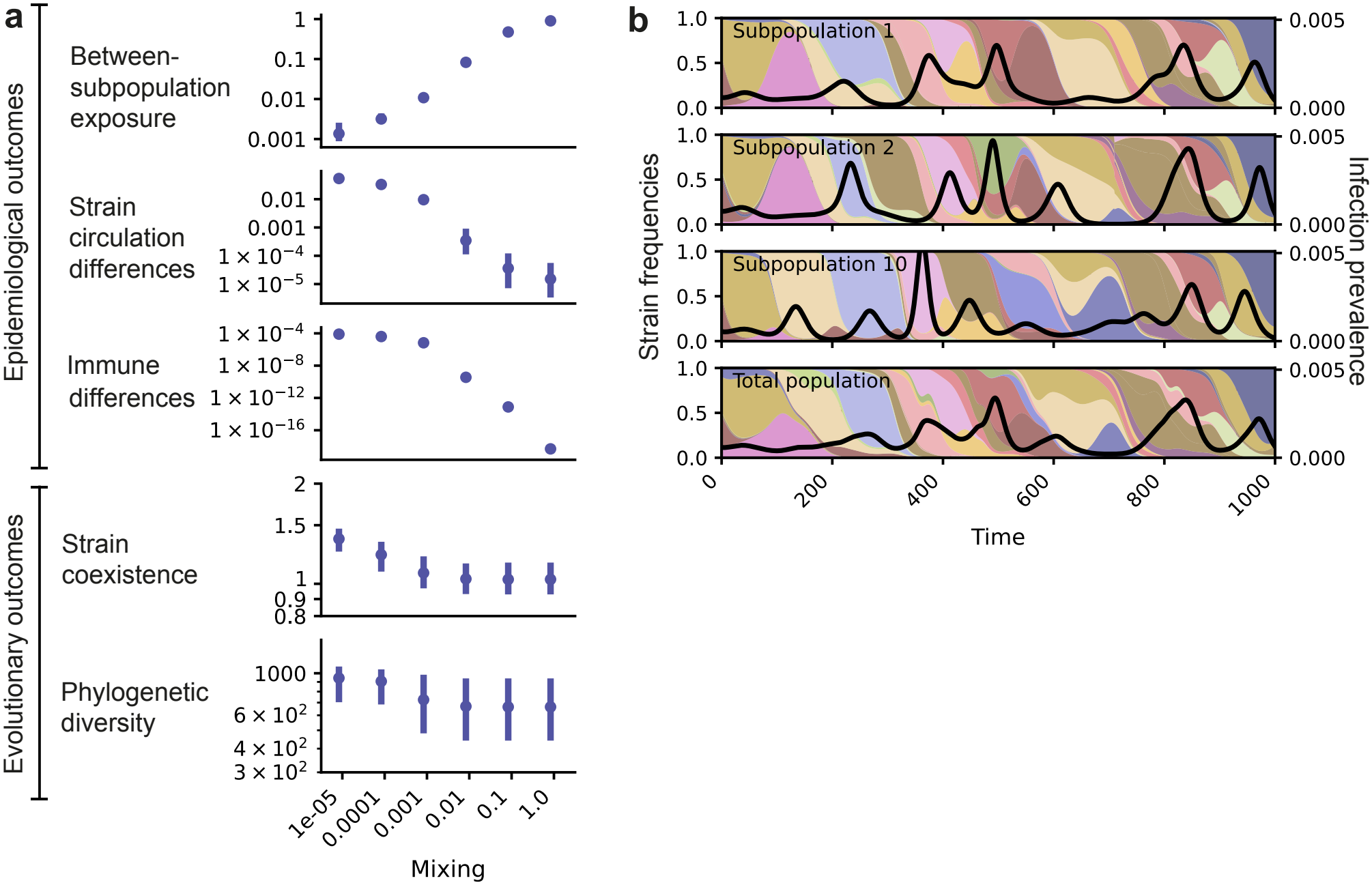
Effects of mixing between subpopulations on the epidemiological and evolutionary dynamics of the model. **a** Model outcomes across different levels of mixing between subpopulations, quantified by the mixing parameter *c*. From top to bottom: proportion of exposure originating from other subpopulations; differences in strain circulation between subpopulations; immune differences between subpopulations; strain coexistence, measured by Shannon entropy; and phylogenetic diversity, measured by the average pairwise *T*_MRCA_. Outcomes were averaged over a 3000*δ*^*−*1^ simulation period. Points show the median and error bars the interdecile range across 100 stochastic simulations. **b** Example dynamics for *c* = 10^*−*4^. Colored areas show the frequencies of circulating strains in each subpopulation (left y-axis), and the black line shows infection prevalence (right y-axis) in that subpopulation. Colors identify the same strains across subpopulations/panels.

Reduced mixing delayed the invasion of strains into other subpopulations and slowed their spread through the overall population. Consequently, rather than generating a single epidemic peak, individual strains produced successive peaks as they spread from one subpopulation to another (Figure 2b). At sufficiently low mixing rates, this delayed spread resulted in differences in the relative prevalence of strains across subpopulations. These differences in strain circulation were accompanied by increasing immune differences between subpopulations, quantified from differences in their strain-specific susceptibility profiles.

Despite these epidemiological and immune differences, evolutionary dynamics changed appreciably only when mixing between subpopulations was extremely limited (Figure 2a, *c ≤* 10^*−*5^). Under these conditions, strain coexistence and phylogenetic diversity, measured by the average pairwise *T*_MRCA_, increased. Thus, limited mixing readily generated differences in strain circulation and immune histories, but substantially altered long-term evolutionary dynamics only under strong population separation.

### Aging increases phylogenetic diversity only in combination with limited mixing

We next investigated aging as a mechanism for generating immune differences between subpopulations and thereby altering evolutionary dynamics (Figure 3). Because immune histories accumulate over time, younger age groups had higher susceptibility and consequently experienced more infections than older age groups (Supplement Figure 7). The magnitude of these age-specific differences was strongly influenced by the breadth of cross-immunity.

**Figure 3.**
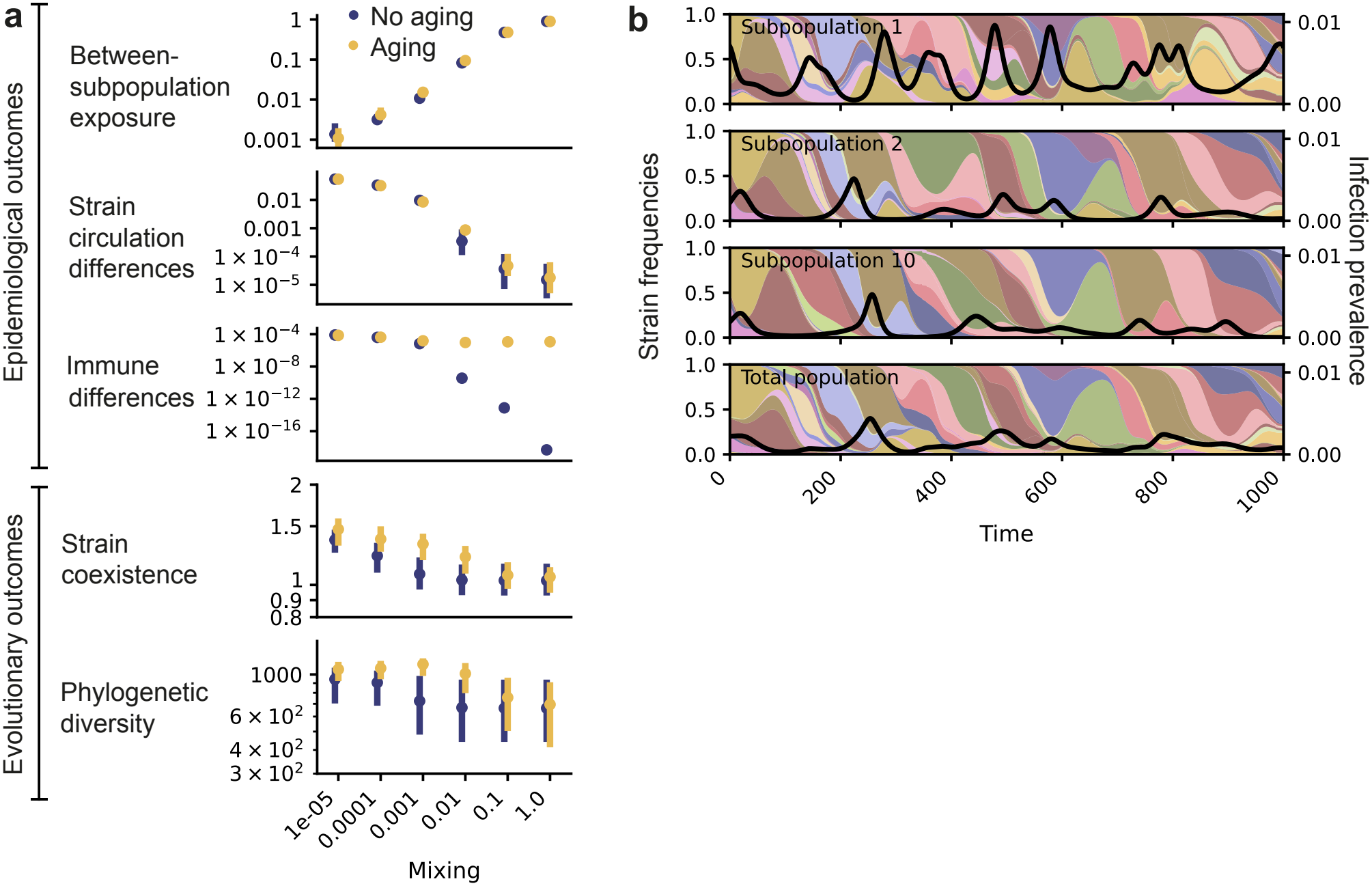
Effects of aging and mixing on the epidemiological and evolutionary dynamics of the model. **a** Model outcomes with and without aging, at different mixing rates. From top to bottom: differences in between-subpopulation exposure; differences in strain circulation between age groups; immune differences between age groups; strain coexistence, measured by Shannon entropy; and phylogenetic diversity, measured by the average pairwise *T*_MRCA_. Outcomes were averaged over a 3000*δ*^*−*1^ simulation period. Points show the median and error bars the interdecile range across 100 stochastic simulations. **b** Example dynamics for *c* = 10^*−*4^. Colored areas show the frequencies of circulating strains in each subpopulation (left y-axis), and the black line shows infection prevalence (right y-axis) in that subpopulation. Colors identify the same strains across subpopulations/panels.

Besides generating differences in overall susceptibility between age groups, aging produced strain-specific immune differences (Figure 3a). However, the strains circulating in different age groups remained similar, as reflected by small between-age-group differences in strain-specific infection patterns. Aging alone therefore had little effect on strain coexistence or phylogenetic diversity.

The effect of aging became more pronounced when combined with limited mixing. Whereas mixing alone altered evolutionary dynamics only at very low mixing rates, the addition of aging increased phylogenetic diversity already at *c ≤* 0.01. This effect was particularly apparent in the youngest age group, which experienced higher infection prevalence and greater strain coexistence than older age groups (Figure 3b for *c* = 10^*−*4^). These dynamics suggest that, under limited mixing, the highly susceptible youngest age group may facilitate the establishment of newly emerging strains while also allowing strains that have been replaced in older age groups to persist for longer. Thus, although age-related differences in immunity alone had little effect on evolutionary dynamics, age structure enhanced the diversification observed when transmission between subpopulations was limited.

### Stochastic immune variation promotes strain replacement rather than coexistence

Finally, we examined whether stochastic immune differences of newly emerging strains could alter evolutionary dynamics (Figure 4). Increasing the magnitude of stochastic variation, as quantified by *σ*, generated greater immune differences between subpopulations. Contrary to our hypothesis, however, these differences did not promote the persistence of divergent strains. Instead, increasing stochastic variation reduced strain coexistence and phylogenetic diversity and resulted in more frequent lineage fixation, shifting the evolutionary dynamics towards successive lineage replacement.

**Figure 4.**
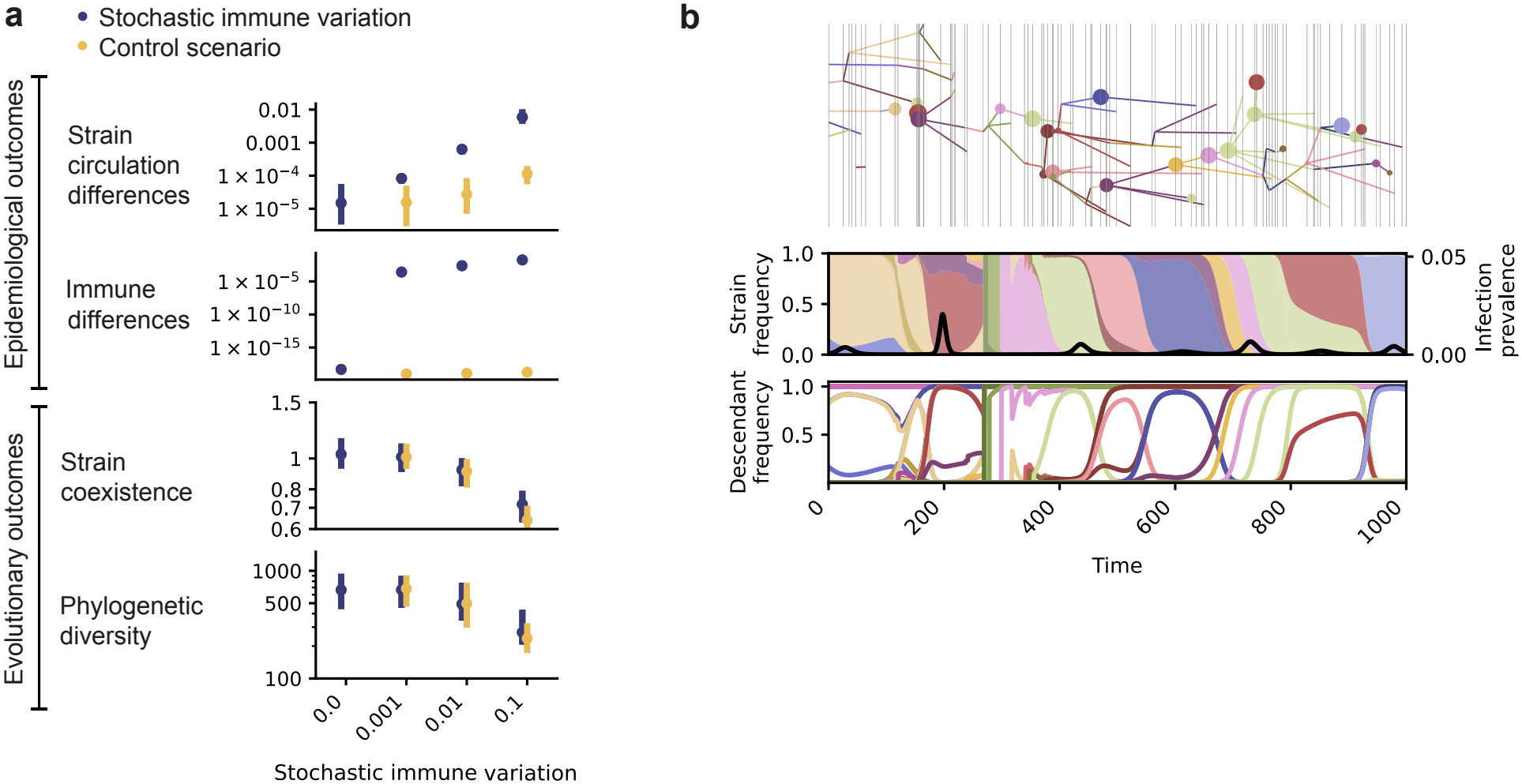
Effects of stochastic immune variation on the epidemiological and evolutionary dynamics of the model. **a** Model outcomes across increasing values of *σ* for stochastic immune variation and the control scenario without between-subpopulation variation. Stochastic immune variation generates both immune differences between subpopulations and variation in the overall susceptibility to emerging strains, whereas the control scenario retains only the latter effect. From top to bottom: differences in strain circulation between subpopulations; immune differences between subpopulations; strain coexistence, measured by Shannon entropy; and phylogenetic diversity, measured by the average pairwise *T*_MRCA_. Outcomes were averaged over a 3000*δ*^*−*1^ simulation period. Points show the median and error bars the interdecile range across 100 stochastic simulations. **b** Example evolutionary and epidemiological dynamics for *σ* = 0.1. The phylogenetic tree shows the emergence and relationships of circulating strains, with node size proportional to cumulative strain prevalence. Colored areas show relative strain frequencies, the black line shows overall infection prevalence, and the final panel shows the frequency of each strain together with all of its descendants. Colors identify the same strains across panels.

This effect could arise not only from immune differences between subpopulations, but also because stochastic variation changed the total number of susceptible hosts available to each newly emerging strain. To distinguish these mechanisms, we considered a control scenario in which stochastic variation altered the total susceptibility to newly emerging strains while preserving the relative susceptibility profiles across subpopulations. This scenario therefore retained variation in the overall susceptible population available to different emerging strains without introducing additional immune differences between subpopulations.

The control scenario produced similar reductions in strain coexistence and phylogenetic diversity and a similar increase in lineage fixation. Thus, the shift towards strain replacement was primarily associated with variation in the overall susceptible population available to newly emerging strains, rather than with additional immune differences between subpopulations. This is consistent with emerging strains that encounter larger susceptible populations having a greater opportunity to spread and replace circulating strains (Figure 4b).

### Evolutionary dynamics depend on the mechanism of strain emergence

We next investigated whether the evolutionary effects of immune differences depended on how novel strains emerged in the model. In the analyses above, the parent strain and subpopulation in which a novel strain emerged were selected uniformly at random, independently of their current prevalence.

We compared these dynamics with a prevalence-weighted strain-emergence mechanism, in which the probability that a novel strain descended from a particular strain in a particular subpopulation was proportional to its relative prevalence. Coupling strain emergence to prevalence changed the baseline evolutionary dynamics. Closely related strains tended to experience similar susceptibility profiles and spread under similar epidemiological conditions, producing pronounced periodic fluctuations in infection prevalence (Supplement Figure 8b). Phylogenetic diversity was also lower than under uniform strain emergence, with the median value of the average pairwise *T*_MRCA_ decreasing from approximately 729 *δ*^*−*1^ to 263 *δ*^*−*1^, and this difference may further increase under longer simulation times. Under uniform strain emergence, even rare strains had the same opportunity as prevalent strains to generate descendants. This increased branching and reduced lineage fixation, as the continual introduction of descendants from different circulating lineages repeatedly disrupted the fixation of prevalent lineages. Despite these differences in baseline evolutionary dynamics, limited mixing, especially in combination with aging, increased phylogenetic diversity under both mechanisms, although the magnitude of the effect was slightly stronger under prevalence-weighted strain emergence (Supplement Figure 9).

Thus, the qualitative relationship between population structure and phylogenetic diversification was robust to the mechanism of strain emergence, but the magnitude of this relationship did depend on the mechanism of strain emergence.

### Sensitivity analyses

We assessed the robustness of the model dynamics to the frequency of strain emergence and the maximum number of co-circulating strains. Decreasing the frequency of strain emergence from *t*_*e*_ = 10 *δ*^*−*1^ to *t*_*e*_ = 100 *δ*^*−*1^ increased strain fixation and resulted in a less branched phylogeny (Supplement Figure 8c). Thus, the rate at which antigenically novel strains were introduced influenced the overall degree of phylogenetic diversification.

We also varied the maximum number of co-circulating strains, *N* , to determine whether the imposed limit constrained the evolutionary dynamics (Supplement Figure 10). We selected *N* = 25 for the main analyses because increasing this limit further had negligible effects on the outcome measures across the parameter ranges considered.

## Discussion

We investigated whether immune differences between host subpopulations alter pathogen antigenic evolution. Using a computationally tractable evolutionary-epidemiological model based on a status-based multi-strain framework [19–22], we considered immune differences generated by mixing, aging, and stochastic immune variation. Overall, our results show that immune heterogeneity does not necessarily promote antigenic diversification. Instead, its evolutionary effects depend on how immune differences are generated and maintained.

Limited mixing generated differences in strain circulation and, consequently, strain-specific immune differences between subpopulations. However, these differences appreciably affected evolutionary dynamics only when mixing was extremely limited (*c ≤* 10^*−*5^). Relating this threshold directly to empirical populations is difficult because *c* represents a simplified ratio of between-to within-subpopulation contacts, whereas real populations have heterogeneous and asymmetric contact and mobility patterns [5, 25, 26]. Nevertheless, the very small values of mixing required in our model suggest that population structure alone substantially increases strain coexistence and phylogenetic diversity only when transmission between subpopulations is strongly restricted. This finding is consistent with previous modeling work showing that spatial separation can promote strain coexistence [27] and with experimental evidence that spatial structure can promote strain diversity [28].

Aging generated a different form of immune heterogeneity. Because immune histories accumulated with age, younger age groups remained more susceptible and experienced more infections, particularly when cross-immunity was broad, reproducing epidemiological patterns observed for infections such as respiratory syncytial virus and measles [29, 30]. Under the uniform strain-emergence mechanism, however, aging alone generated relatively little strain-specific differences in immunity and consequently had little effect on strain coexistence or phylogenetic diversity. Its effects became more pronounced when combined with limited mixing, with phylogenetic diversity increasing at mixing levels for which mixing alone had little evolutionary effect. One possible explanation is that the highly susceptible youngest age group provides an epidemiological niche in which newly emerging strains can establish and existing lineages can persist for longer when transmission between age groups is restricted.

This mechanism is unlikely to operate strongly through age structure alone in realistic populations. Empirical contact studies show substantial assortative mixing by age, but also frequent contacts between age groups [25, 26]. In our model, by contrast, aging substantially increased phylogenetic diversity only when transmission between subpopulations was strongly restricted. Limited mixing and age-related differences in immune histories alone are therefore unlikely to explain the age-specific antigenic selection reported for influenza [8]. More generally, however, our results suggest that highly susceptible subpopulations that are partially separated from the rest of the population may provide epidemiological niches that facilitate the establishment and persistence of distinct pathogen lineages.

Stochastic immune variation produced a qualitatively different and initially counterintuitive effect. Rather than promoting strain coexistence and phylogenetic diversity, increasing stochastic variation favored lineage fixation and successive strain replacement. The control scenario helped distinguish the mechanism underlying this result. Stochastic variation generated not only immune differences between subpopulations but also variation among emerging strains in the overall susceptible population available to them. Retaining the latter variation while removing the additional between-subpopulation immune differences produced similar evolutionary dynamics. This suggests that the shift towards lineage replacement was driven primarily by variation in the overall susceptible population encountered by emerging strains.

Strains that encounter a relatively large susceptible population have a greater opportunity to spread, creating asymmetries in epidemiological success that favor replacement rather than coexistence. Mechanistically, this resembles how the strength and breadth of cross-immunity determine whether competing strains coexist or replace one another [31, 32].

Furthermore, assumptions governing the emergence of novel strains influence the evolutionary dynamics in our model. The qualitative effects of mixing, aging, and stochastic immune variation were similar under different emergence mechanisms, but their magnitude differed. When strain emergence was prevalence-weighted, epidemiological success increased the opportunity for a lineage to generate antigenically novel descendants, which may strengthen the evolutionary consequences of differences in transmission and susceptibility between subpopulations.

Taken together, these findings suggest that the evolutionary consequences of immune heterogeneity depend on the type of heterogeneity generated. Differences in overall susceptibility or transient stochastic variation are not sufficient to promote diversification and may instead favor replacement. In contrast, persistent differences in strain-specific susceptibility between partially separated host subpopulations can create distinct epidemiological and immunological environments in which different lineages persist. However, in our model, substantial effects generally required strong population separation or an interaction between population structure and other sources of heterogeneity. This is consistent with previous modeling showing that immune heterogeneity combined with assortative mixing can increase strain emergence, although the magnitude of this effect may be small relative to other determinants of pathogen evolution [14]. The framework could be extended to investigate whether other processes strengthen or weaken these effects, including heterogeneity in infectivity, seasonality, and contact rates [12], waning immunity [33], and immune imprinting or original antigenic sin [10, 24].

Beyond these biological findings, our results highlight several challenges inherent to evolutionary-epidemiological modeling. A first challenge is representing large numbers of antigenically related strains while retaining epidemiologically meaningful immune interactions. To remain computationally tractable, multi-strain models necessarily make simplifying assumptions. The status-based framework used here scales linearly with the number of strains but assumes independence among co-infecting strains and does not represent direct interactions between them [34]. History-based models instead track individual infection histories, but their state space grows rapidly with the number of strains and they therefore commonly require other simplifications, such as assuming that cross-immunity is determined primarily by the most recent infection [35–37]. Other approaches reduce the dimensionality of immune histories through age structure [38] or restrict antigenic evolution to occur between epidemic seasons [24]. Different modeling frameworks therefore make different trade-offs between epidemiological detail, immune history, evolutionary dynamics, and computational tractability.

A second challenge is representing the emergence of novel strains. Our results were quantitatively sensitive to whether novel strains emerged uniformly across circulating strains and subpopulations or in proportion to their prevalence, as well as to the frequency of strain emergence (see also [39]). These assumptions determine how strongly epidemiological success feeds back into evolutionary opportunity. Uniform emergence gives rare and prevalent lineages similar opportunities to generate descendants, whereas prevalence-weighted emergence favors lineages responsible for more infections. Neither representation captures the complete biological process of mutation and establishment. A natural extension would be to link the generation of novel variants directly to the number of infections while allowing their antigenic effects to vary, thereby distinguishing the generation of mutations from their subsequent epidemiological establishment.

A third challenge is determining which model behaviors can be meaningfully compared with empirical evolutionary dynamics. Summary measures calculated from simulations can depend on the simulation duration, burn-in period, temporal autocorrelation, and stochastic variation within and between simulations. Empirical measures derived from sequencing data, strain prevalence, or immune profiles are subject to different limitations, including incomplete sampling and measurement error. Direct comparisons between simulated and observed evolutionary patterns may therefore depend strongly on the outcome chosen. Developing robust and interpretable measures that can be calculated consistently from both model output and empirical data will be important for evaluating alternative model assumptions and determining which mechanisms reproduce observed pathogen evolutionary dynamics.

Our study has several limitations. The model deliberately represents antigenic evolution in a simplified manner. Each novel strain differs from its parent by a single antigenic step, antigenic distance is defined on a tree without recurrent or back mutations, and cross-immunity is determined solely by this distance. Strain emergence is imposed as a stochastic process rather than arising explicitly from mutation within infected hosts, and extinction is evaluated at strain-emergence events. The model also excludes several processes that may generate or modify immune heterogeneity in real populations, including waning immunity, vaccination, seasonality, variation in transmission rates, and more complex individual immune histories. These simplifications allowed us to isolate the mechanisms considered here, but they limit direct quantitative interpretation for any particular pathogen. The mixing thresholds identified in our simulations should therefore be interpreted as properties of the model and parameter regime rather than universal thresholds at which population structure begins to influence antigenic evolution.

In conclusion, immune heterogeneity can shape antigenic evolution, but its presence alone does not necessarily promote pathogen diversification. In our model, diversification was favored when population structure generated sufficiently persistent strain-specific differences between host subpopulations, whereas stochastic variation in susceptibility instead promoted strain replacement. These findings highlight the potential role of host population structure in shaping antigenic evolution. However, further progress will require models whose assumptions about immune interactions and strain emergence can be systematically evaluated against robust measures derived from empirical pathogen and immunity data.

## Data Availability

The model code is available at https://github.com/MyrtheW/immune_heterogeneity_evolution

## Data Accessibility

The model code is available at https://github.com/MyrtheW/immune_heterogeneity_evolution

## Authors’ Contributions

G.R. acquired funding. G.R. and R.A.N. supervised the study. M.S.W, G.R., and R.A.N. developed the mathematical model and the scenarios. M.S.W carried out data analysis, wrote computer code, conducted and planned simulations, and prepared figures. M.S.W. and G.R. wrote the manuscript. R.A.N. reviewed the manuscript.

## References

1 A. K. Oraby et al., “A single amino acid mutation alters multiple neutralization epitopes in the respiratory syncytial virus fusion glycoprotein”, npj Viruses 3, 1–14 (2025).

2 A. Z. Mykytyn, R. A. Fouchier, and B. L. Haagmans, “Antigenic evolution of SARS coronavirus 2”, Current Opinion in Virology 62, 10.1016/j.coviro.2023.101349 (2023).

3 K. E. Kistler and T. Bedford, “An atlas of continuous adaptive evolution in endemic human viruses”, Cell Host & Microbe 31, 1898–1909.e3 (2023).

4 A. X. Han, S. P. de Jong, and C. A. Russell, “Co-evolution of immunity and seasonal influenza viruses”, Nature Reviews Microbiology 21, 805–817 (2023).

5 T. Bedford et al., “Global circulation patterns of seasonal influenza viruses vary with antigenic drift”, Nature 523, 217–220 (2015).

6 C. A. Russell et al., “The global circulation of seasonal influenza A (H3N2) viruses”, Science 320, 340–346 (2008).

7 M. R. Islam, M. N. Hoque, M. S. Rahman, A. S. U. Alam, M. Akther, J. A. Puspo, S. Akter, M. Sultana, K. A. Crandall, and M. A. Hossain, “Genome-wide analysis of SARS-CoV-2 virus strains circulating worldwide implicates heterogeneity”, Scientific Reports 10, 14004–(2020).

8 F. C. Welsh et al., “Age-dependent heterogeneity in the antigenic effects of mutations to influenza hemagglutinin”, bioRxiv, 10.1101/2023.12.12.571235 (2023).

9 C. A. Dendrou, J. Petersen, J. Rossjohn, and L. Fugger, “HLA variation and disease”, Nature Reviews Immunology 18, 325–339 (2018).

10 S. Cobey and S. E. Hensley, “Immune history and influenza virus susceptibility”, Current Opinion in Virology 22, 105–111 (2017).

11 D. J. Smith, A. S. Lapedes, J. C. D. Jong, T. M. Bestebroer, G. F. Rimmelzwaan, A. D. Osterhaus, and R. A. Fouchier, “Mapping the antigenic and genetic evolution of influenza virus”, Science 305, 371–376 (2004).

12 A. J. Kucharski and J. R. Gog, “Age profile of immunity to influenza: effect of original antigenic sin”, Theoretical Population Biology 81, 102–112 (2012).

13 H. Chabas, S. Lion, A. Nicot, S. Meaden, S. van Houte, S. Moineau, L. M. Wahl, E. R. Westra, and S. Gandon, “Evolutionary emergence of infectious diseases in heterogeneous host populations”, PLOS Biology 16, e2006738 (2018).

14 A. Yates, R. Antia, and R. R. Regoes, “How do pathogen evolution and host heterogeneity interact in disease emergence?”, Proceedings of the Royal Society B: Biological Sciences 273, 3075 (2006).

15 A. E. Fleming-Davies, V. Dukic, V. Andreasen, and G. Dwyer, “Effects of host heterogeneity on pathogen diversity and evolution”, Ecology Letters 18, 1252–1261 (2015).

16 G. Rozhnova, A. Nunes, and A. J. McKane, “Impact of commuting on disease persistence in heterogeneous metapopulations”, Ecological Complexity 19, 124–129 (2014).

17 T. J. Hagenaars, C. A. Donnelly, and N. M. Ferguson, “Spatial heterogeneity and the persistence of infectious diseases”, Journal of Theoretical Biology 229, 349–359 (2004).

18 M. J. Keeling and P. Rohani, “Estimating spatial coupling in epidemiological systems: A mechanistic approach”, Ecology Letters 5, 20–29 (2002).

19 J. R. Gog and B. T. Grenfell, “Dynamics and selection of many-strain pathogens”, Proceedings of the National Academy of Sciences 99, 17209–17214 (2002).

20 J. R. Gog and J. Swinton, “A status-based approach to multiple strain dynamics”, Journal of Mathematical Biology 44, 169–184 (2002).

21 B. Grenfell and J. Gog, “Pathogen strains: no joke”, Trends in Ecology & Evolution 16, 272 (2001).

22 P. Barrat-Charlaix and R. A. Neher, “Eco-evolutionary dynamics of adapting pathogens and host immunity”, eLife 13, RP97350 (2024).

23 L. Yan, R. A. Neher, and B. I. Shraiman, “Phylodynamic theory of persistence, extinction and speciation of rapidly adapting pathogens”, eLife 8, e44205 (2019).

24 A. J. Kucharski and J. R. Gog, “The role of social contacts and original antigenic sin in shaping the age pattern of immunity to seasonal influenza”, PLOS Computational Biology 8, e1002741 (2012).

25 J. Mossong et al., “Social Contacts and Mixing Patterns Relevant to the Spread of Infectious Diseases”, PLOS Medicine 5, e74 (2008).

26 D. Mistry et al., “Inferring high-resolution human mixing patterns for disease modeling”, Nature Communications 12, 323–(2021).

27 M. Lipsitch, C. Colijn, T. Cohen, W. P. Hanage, and C. Fraser, “No coexistence for free: Neutral null models for multistrain pathogens”, Epidemics 1, 2 (2008).

28 E. Visher et al., “Spatially structured host genetic diversity leads to the evolution of local specialization”, Evolution 80, 800–811 (2026).

29 A. C. Langedijk and L. J. Bont, “Respiratory syncytial virus infection and novel interventions”, Nature Reviews Microbiology 21, 734–749 (2023).

30 J. Singh and K. K. Datta, “Epidemiological Considerations of the Age Distribution of Measles in India: a Review”, Journal of Tropical Pediatrics 43, 111–115 (1997).

31 S. Gupta, N. Ferguson, and R. Anderson, “Chaos, persistence, and evolution of strain structure in antigenically diverse infectious agents”, Science 280, 912–915 (1998).

32 N. M. Ferguson, A. P. Galvani, and R. M. Bush, “Ecological and immunological determinants of influenza evolution”, Nature 422, 428–433 (2003).

33 S. Ranjeva, R. Subramanian, V. J. Fang, G. M. Leung, D. K. Ip, R. A. Perera, J. S. Peiris, B. J. Cowling, and S. Cobey, “Age-specific differences in the dynamics of protective immunity to influenza”, Nature Communications 10, 1–11 (2019).

34 E. Gjini, C. Valente, R. Sá-Leão, and M. G. M. Gomes, “How direct competition shapes coexistence and vaccine effects in multi-strain pathogen systems”, Journal of Theoretical Biology 388, 50–60 (2016).

35 J. Lin, V. Andreasen, R. Casagrandi, and S. A. Levin, “Traveling waves in a model of influenza A drift”, Journal of Theoretical Biology 222, 437–445 (2003).

36 I. M. Rouzine and G. Rozhnova, “Antigenic evolution of viruses in host populations”, PLOS Pathogens 14, e1007291 (2018).

37 M. S. Willemsen and G. Rozhnova, “Impact of vaccination on the speed of antigenic evolution”, medRxiv, 2026.03.10.26347605 (2026).

38 A. J. Kucharski, V. Andreasen, and J. R. Gog, “Capturing the dynamics of pathogens with many strains”, Journal of Mathematical Biology 72, 1 (2015).

39 P. Minayev and N. Ferguson, “Incorporating demographic stochasticity into multi-strain epidemic models: application to influenza A”, Journal of the Royal Society Interface 6, 989–996 (2009).

