## Supplement for "Impact of immune heterogeneity on antigenic evolution"

### Supplementary Material


royalsocietypublishing.org/journal/rsif J R Soc Interface 0000000

### 1. Supplementary Methods

#### (a) Model implementation

The implementation is developed in Python 3.13.2 and relies on the following packages: Pandas 2.3.2, SciPy 1.15.2, PyYAML 6.0.2, and NumPy 2.2.3. Numpy was utilized to accelerate the calculation of the ODEs through vectorized operations. The ODEs were solved using the 'solve_ivp' function with the Runge-Kutta method of order 5 (RK45).

##### (i) Computation of $T_{\mathrm{MRCA}}$

To efficiently compute the time elapsed since the most recent common ancestor of strains $a$ and $b$ emerged, $T_{\mathrm{MRCA}}(a,b,t)$, we track the emergence time of the most recent common ancestor, $T'_{\mathrm{MRCA}}(a,b)$, for all pairs of co-circulating strains. When a novel strain $a$ emerges, we set $T'_{\mathrm{MRCA}}(a,a)$ to its own emergence time and inherit $T'_{\mathrm{MRCA}}(a,b)$ for all other strains $b$ from its parent $p$, such that $T'_{\mathrm{MRCA}}(a,b) = T'_{\mathrm{MRCA}}(p,b)$. Finally, we calculate phylogenetic diversity by subtracting the weighted average emergence time from the current time.

#### (b) Approximating susceptibility to novel strains

When a novel strain emerges, certain individuals will be immune through prior exposure to antigenically related strains. When these full exposure histories are not available, determining the proportion of susceptible individuals $S_i^a$ to the novel strain $a$ is not trivial. We derived an approximation for $S_i^a$ by integrating $\dot{S}_i^a$. Here we assume that population turnover, $\gamma < 1/t_e, \gamma \ll \delta$ has limited impact on these dynamics:

$$S_i^a(t) = S_i^a(0)\exp\left(-\alpha \sum_{j=1}^{M} C_{ij} \sum_{b=1}^{N} K^{ab} \int_0^t I_j^b(t), dt\right)$$

In this equation, we substitute $S_i^a(0) = 1$ and $K^{ab} = e^{-\frac{D(a,b)}{d}} = e^{-\frac{D(a,p)+D(p,b)}{d}} = K^{ap}K^{pb}$ (since the parent strain $p$ lies on the path between $a$ and any other strain $b$), we can rewrite this as:

$$\begin{aligned}
S_i^a(t) &= \exp\left(-\alpha \sum_{j=1}^{M} C_{ij} \sum_{b=1}^{N} K^{ap}K^{ab} \int_0^t I_j^b(t)dt\right)\\
&= \exp\left(K^{ap}(-\alpha \sum_{b=1}^{N}\sum_{j=1}^{M} C_{ij}K^{pb} \int_0^t I_j^b(t)dt)\right)\\
&= \exp\left(K^{ap}\ln\left(S_i^p(t)\right)\right)\\
&= S_i^p(t)^{K^{ap}}
\end{aligned}$$

### 2. Supplementary Algorithms

**Algorithm 1** Introducing a novel strain

1: Jointly select a parent strain $p$ and subpopulation $i$, at random or weighted by prevalence $I_i^p(t)$.
2: If the maximum number of strains $N$ is reached: Remove the strain $c$ with the lowest overall prevalence $\sum_{i=1}^{M} I_i^c(t)$ .
3: Compute distance $D(a,b) = D(p,b) + 1$ for each circulating strain $b$.
4: Compute cross-immunities $K^{ab}$
5: Approximate the individuals susceptible to strain $a$, $S_i^a(t) = S_i^p(t)^{K^{ap}}$
6: Initialize infections $I_i^a(t) = 10^{-6}$ for strain $a$ in subpopulation $i$.

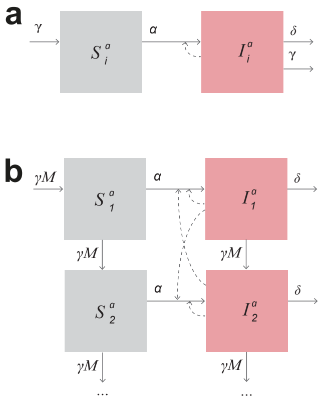

**Figure 5. Model schematic.** Transitions are shown for a single strain. **a** Transitions within a single subpopulation. **b** Incorporation of age structure between subpopulations.

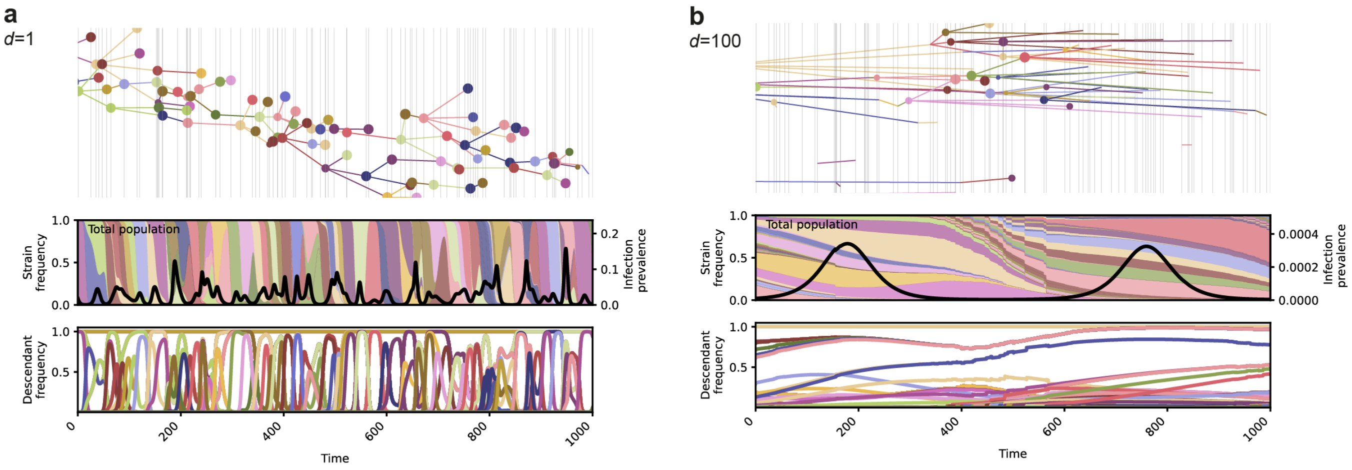


**Figure 6. Effect of cross-immunity breadth on evolutionary dynamics.** Evolutionary dynamics for two alternative values of the cross-immunity breadth, (**a**) $d = 1$ and (**b**) $d = 100$. The upper panels show the phylogenetic tree of circulating strains. The horizontal position of each node indicates the time at which the corresponding strain emerged, while the vertical position is arbitrary. Light grey vertical lines indicate strain-introduction events, and node size is proportional to the cumulative prevalence of the corresponding strain. **b** Relative frequencies of circulating strains (colored areas, left y-axis) and overall infection prevalence (black line, right y-axis). **c** Frequency of each strain together with all of its descendants, illustrating the persistence and fixation of individual lineages. Colors identify the same strains across panels.

### 3. Supplementary Figures

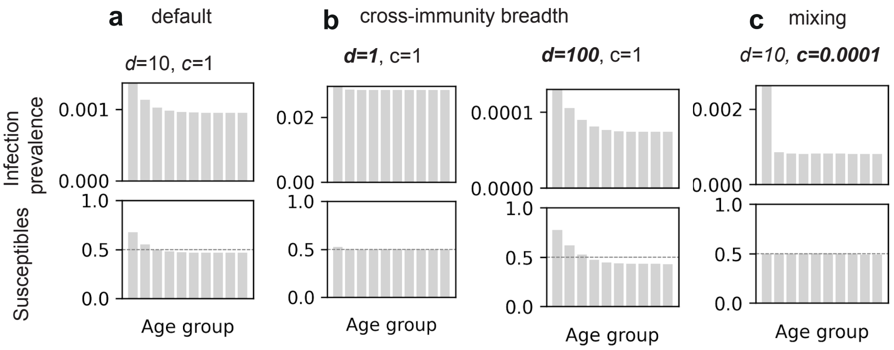


**Figure 7. Effect of cross-immunity breadth $d$ and mixing $c$ on overall (non-strain-specific) differences in infection prevalence and susceptibility.** **a** Age-specific infection prevalence (top) and susceptibility (bottom) under baseline parameters. **b** Effect of cross-immunity breadth ($d = 1$ and $d = 100$), and **c** effect of slow mixing ($c = 0.001$).

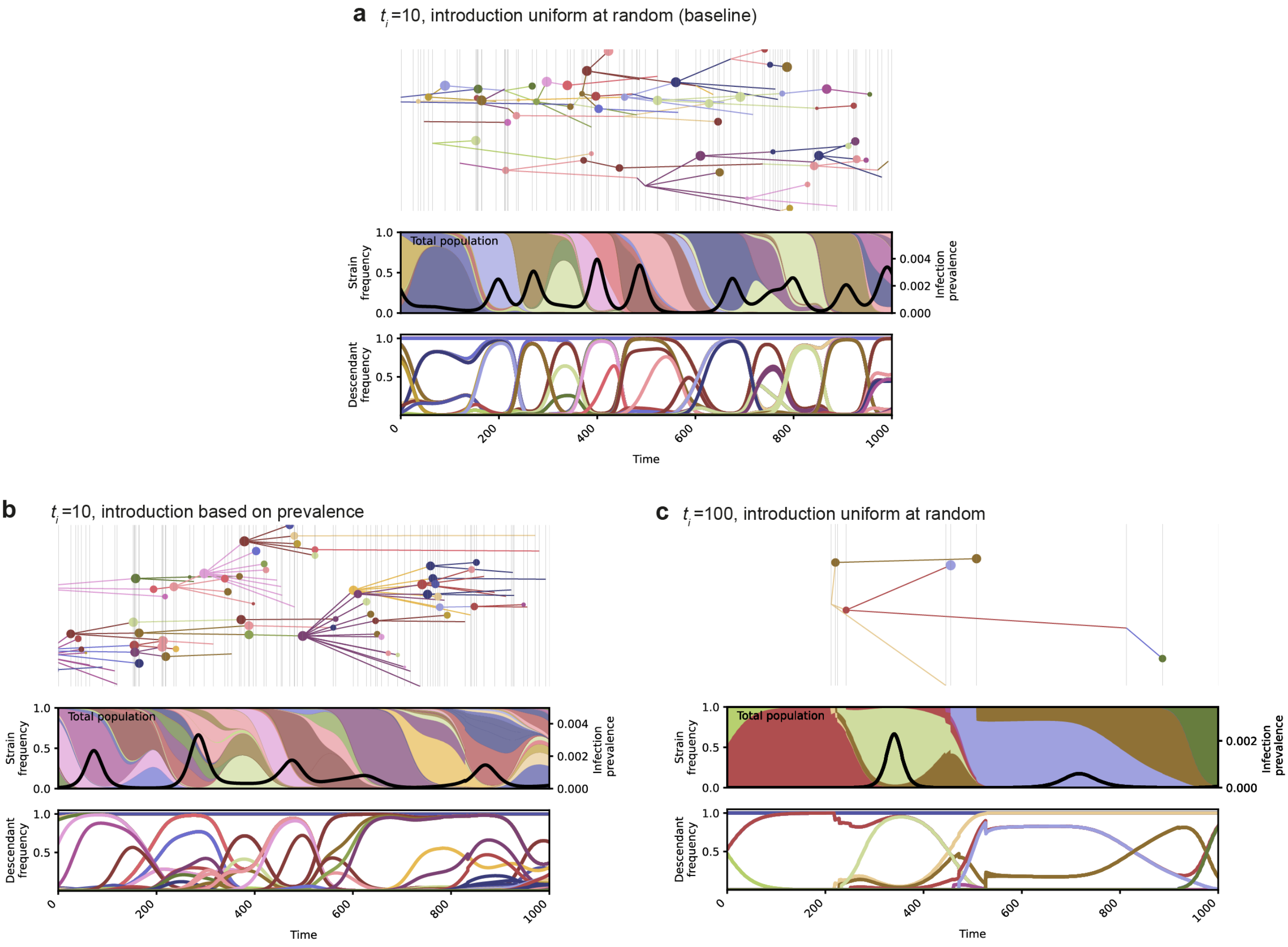


**Figure 8. Evolutionary dynamics under different strain introduction methods.** **a** Baseline scenario: parent strains are selected uniformly at random, with a mean introduction interval of $10\delta^{-1}$. **b** Parent strains are selected in proportion to their prevalence. **c** Mean introduction interval increased from 10 to $100\delta^{-1}$. See Figure 1 for panel descriptions.

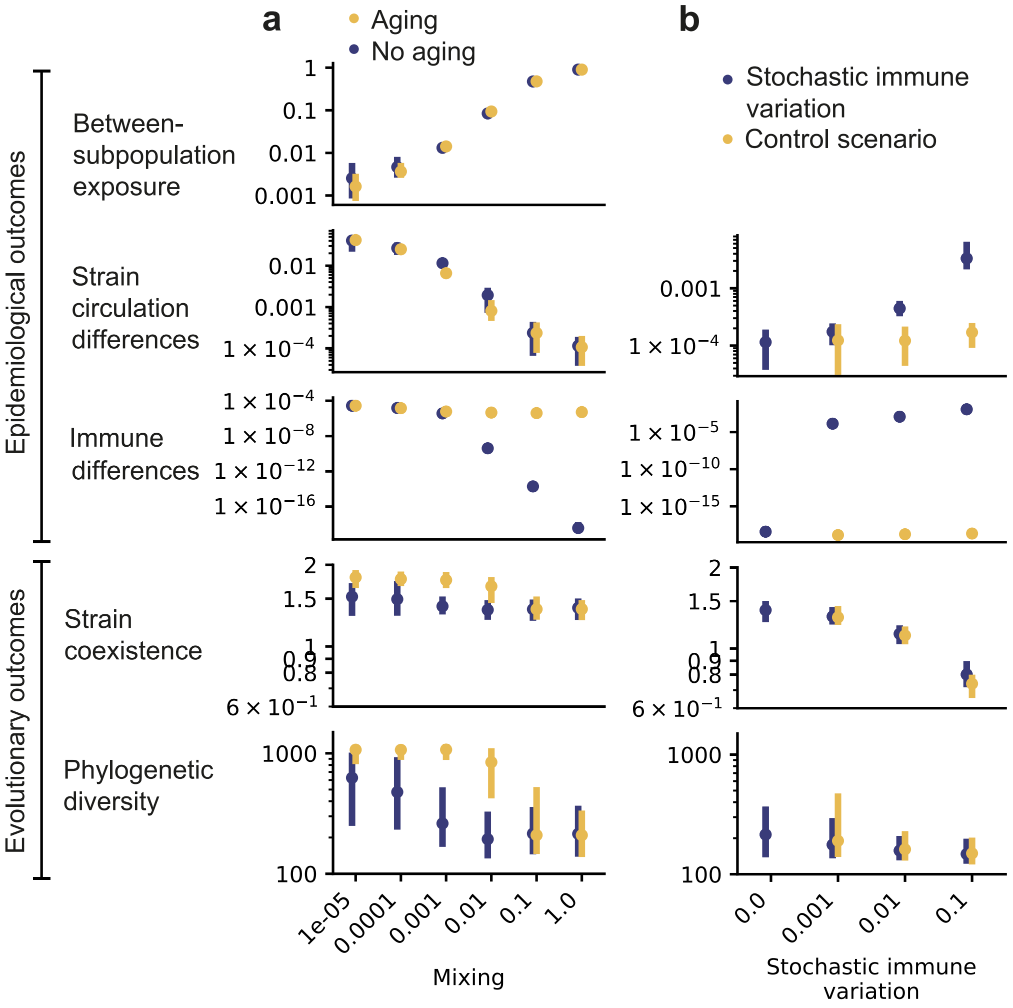


**Figure 9. The effect of immune differences on evolution under prevalence-based introduction of novel strains.** Effect of **a** mixing, **b** aging and **c** stochastic immune variation on (from top to bottom): differences in between-subpopulation exposure; differences in strain circulation between age groups; immune differences between age groups; strain coexistence, measured by Shannon entropy; and phylogenetic diversity, measured by the average pairwise $T_{\mathrm{MRCA}}$. Outcomes were averaged over a $3000\delta^{-1}$ simulation period. Points show the median and error bars the interdecile range across 100 stochastic simulations. **b** example dynamics for $c = 10^{-4}$. Colored areas show the frequencies of circulating strains in each subpopulation (left y-axis), and the black line shows infection prevalence (right y-axis) in that subpopulation. Colors identify the same strains across subpopulations/panels.

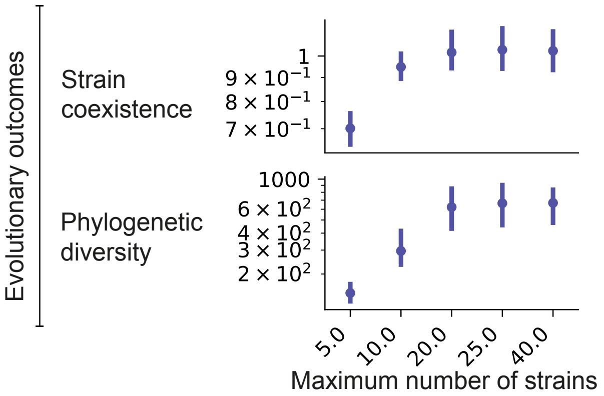


**Figure 10. Effect of maximum number of strains on strain coexistence and phylogenetic diversity.** The strain coexistence was measured by Shannon entropy and the phylogenetic diversity by the average pairwise $T_{\mathrm{MRCA}}$. Outcomes were averaged over a $3000\delta^{-1}$ simulation period. Points show the median and error bars the interdecile range across 100 stochastic simulations.
